# Reducing suicide attempts with a compounded Brief Contact Intervention: a nationwide study of the VigilanS project, France

**DOI:** 10.64898/2026.02.05.26345598

**Authors:** Xavier Xu Wang

## Abstract

**Background:** Suicide prevention has become a global public health priority, and Brief Contact Interventions (BCI) following suicide attempts (SA) are an important tool for preventing suicides. The VigilanS project was designed to generalize compounded BCIs at the entire population level., It involves resource cards, telephone calls, and mailings, following a predefined algorithm. It has been implemented progressively in France, on a region-by-region basis, since 2015.

**Objective:** To evaluate the effectiveness of VigilanS in reducing suicide attempts among patients aged 18 years and older, and to explore potential differences in effectiveness by sex, age, and geographical location.

**Methods:** The study used data from the French national hospitalization database, PMSI-MCO. It included all patients over age 18 who were admitted to general hospitals for suicide attempts, between 2012 and 2022. Time-to-event (“survival”) analysis of a second SA after a first one was performed; patients whose first SA occurred before VigilanS implementation were compared with their after-VigilanS counterparts. Six regions, with implementation occurring between 2015 and 2017, are analyzed here.

**Results:** The differences in distribution of time-to-new-SA among patients before and after VigilanS implementation were statistically significant in all six regions under scope (log-rank test: P<0.0001). The Cox regression analysis revealed that VigilanS was significantly associated with a reduced risk of reattempting suicide in all regions. Age consistently showed a negative association with reattempting suicide.

**Conclusion:** VigilanS is likely effective in reducing suicide attempts among patients aged 18 years and older in France. This suggests that implementing BCIs following SAs in general hospitals at a population-wide level can contribute to reducing suicide rates and provides real-world evidence (RWE).

## Context

Suicide and self-harm are significant concerns in France, accounting for one of the top ten countries with the highest suicide rates in Europe [1]. Several countries in Europe have developed national suicide prevention strategies and large-scale multimodal projects are considered the most effective strategy to adopt [2,3]. In France, it is estimated that between 176,000 and 200,000 suicide attempts per year lead to hospital visits due to their physical or psychological severity[4]. This highlights the importance of implementing prevention projects in general hospitals.

An important aspect of preventing suicide attempts (SA) and suicides is the implementation of Brief Contact Interventions (BCI) following a SA [5–8]. A recent meta-analysis evaluates the effectiveness of BCIs in preventing suicidal deaths, attempts, and ideations among patients with mental health disorders discharged from the hospital, suggesting that BCIs significantly reduced suicidal attempts and ideations within certain time frames; there were no significant findings on suicidal deaths [9]. Another meta-analysis of 14 clinical trials published between January 2000 and May 2019 found that brief acute care interventions were associated with reduced subsequent suicide attempts, and increased adherence to follow-up care, but were not associated with reduced depression symptoms. The study concluded that single in-person encounter may be effective at reducing subsequent suicide attempts and at ensuring that patients adhere to follow-up mental health care [10]. VigilanS is a compounded BCI implemented in entire France regions. It uses resource cards, telephone calls, and mailings following a predefined algorithm [11]. VigilanS is implemented in all structures in a department or region that are likely to attend patients who have attempted suicide [12,13].

In the absence of an ad hoc registry in France, it is difficult to accurately measure the number of SA and to follow their evolution [14,15]. Therefore, the most commonly used measure is hospitalization data, specifically the PMSI system, which provides a standardized and synthetic description of medical activity in healthcare institutions, including hospital stays related to SA[16]. This system comprises four main subsystems, depending on the institution specialization, and includes medicine, surgery, obstetrics and odontology (MCO ; general hospitals), follow-up and rehabilitation care (SSR), psychiatry (Psy), and hospitalization at home (HAD) [17]. In this study, PMSI-MCO was utilized to extract the data of hospitalized patients at the national level.

## Objectives

The main objective of this study is to evaluate the effectiveness of VigilanS in reducing suicide attempts among patients aged 18 years and older, and to explore potential differences in effectiveness by sex, age, and geographical location. This will be accomplished by performing time-to-event analysis of a second SA after a first one, on PMSI-MCO data.

## Methods

### Description of the VigilanS project

VigilanS is a monitoring intervention that follows patients up for six months after they have experienced a SA. The system is organized into surveillance cells, with one cell per geographical area of implementation (former region or department). The health care centers get patients enrolled by sending an encrypted fax to the ad hoc VigilanS cell, or by securing registration on a dedicated computer platform.

When a patient is discharged from the hospital, he is given a resource card with the VigilanS phone number on it, as well as other emergency numbers. From that moment onward, VigilanS is responsible for managing the patient’s intervention and ongoing care, in addition to the regular care provided by the participating medical center, for an initial period of six months [13]. Non-first-time suicide attempters are called between the 10th and 21st day after hospital discharge, to assess their situation and decide on an appropriate action, which may include scheduling an emergency or regular appointment, scheduling another call, sending personalized postcards, or taking no further action. After the six-month follow-up period, all patients are contacted for an end-of-follow-up interview. If necessary, the intervention is extended for an additional 3 or 6 months. Patients who attempt suicide during the follow-up period are reset for another 6 months. In addition to the two scheduled calls at D10-D21 and at 6 months, intermediate calls may be made by VigilanS or patients themselves.

There are no restrictions on including a patient in the project, except that he must be reachable by telephone and mail, since these are the primary means of communication of the system; they must reside in the region or department of the attending center (referred to as “center” hereafter). SA is defined as actively, voluntarily, and intentionally trying to end one’s life, which corresponds to the international definition [18], as well as the definition used in all the previous work related to VigilanS.

Since January 2015, the VigilanS project has been implemented in the Nord-Pas-de-Calais region [19], after a year (2014) of testing with a subset of centers. It has been progressively implemented in five other geographical areas: Basse Normandie (March 2016), the Jura department (April 2016), Languedoc-Roussillon (May-June 2016), Bretagne (June 2016), and Haute Normandie (March 2017). By the end of 2017, more than 10,000 subjects were included in the project, and another 10,000 joined in 2018. Since 2019, VigilanS has been implemented throughout France (see Appendix 1), under the responsibility of the project’s design team (Pr Vaiva, Lille).

### Data processing and statistical analysis

The study population comprises all patients over the age of 18 who have been admitted to general hospitals for suicide attempt. This population is exhaustively recorded in the PMSI-MCO, for the analysis period 2012-2022. Data extractions were carried out from that database; records were filtered on the main diagnoses (From X60 to X84, self-inflicted injury, poisonings and some other consequences of external causes) [20]. No individual-level VigilanS internal database or operational files were accessed; all analyses were exclusively based on PMSI-MCO hospitalization records.

The hospital identification number was used to determine its geographical region; alongside with hospitalization date, it allowed us to determine whether or not VigilanS implementation had started or not. Stays for the same patient were linked together using variables specially designed for this purpose. This shifted the analysis from being based on hospital stays to an analysis based on patients. For patients who have had multiple hospitalizations due to SA during the 2012-2022 period, we used the two first stays only.

We performed survival analysis (time-to-event analysis) to estimate the probability of a SA reattempt over time [21]; we used the Kaplan-Meier estimate, log-rank test and Cox proportional hazards model. We analyzed SA reattempts between January 1, 2012 and December 31, 2022, and defined, for each region, a pre-VigilanS group and a post-VigilanS group, based on the date of each patient’s first SA relative to the VigilanS inception date. The starting point of each observation is a first SA in the 2012-2022 window, and the ending point is the first reoccurrence in the same window; subsequent reoccurrences are not considered. When the first SA occurs before VigilanS, the patient is classified in the “pre” group; and vice versa for the “post” group; each region has its own before/after separation date (month and year), since inception date is region-specific. For the NPC, we excluded 2014, that corresponds to the aforementioned testing year; “pre” therefore corresponds to a first TS in 2012 or 2013, and “post” corresponds to a first TS in 2015 or subsequent years. The “pre” observation period (VigilanS = 0, red curves) is longer, since it can go up to 11 years (extreme case: 1st TS on 01/01/2012, 1st reiteration on 31/12/2022), whereas the “post” period (VigilanS = 1, turquoise curves) only starts in 2015 or 2016 or 2017 depending on the case (beginning or middle of the year), while ending on 31/12/2022.

In the cox proportional hazards model, the dependent variable (outcome) is the suicide reattempt. The independent variables include VigilanS (as defined above), gender, and age. Separate modeling was conducted for each region, because of different VigilanS implementation start dates, specific healthcare infrastructures, regional manaagement of healthcare in France, and specific adaptations of VigilanS design.

All statistical analysis was done using the R statistical software version 2022.07.1.

## Results

### Number of patients hospitalized annually for SA

A total of 204,875 patients (> 18 years old) were hospitalized for suicidal attempts (SA) between 2012 and 2022 in the six regions under scope. Table 1gives the number of SA cases across regions and calendar years. The region with the highest number of SA cases was Nord-Pas-de-Calais (population 4.06 million), with a total of 76,156 patients hospitalized for SA. This indicates a significant burden of SA in this region. On the other hand, the Jura department (population 258, □ 624) had the lowest number of SA cases, with a total of 3,779 patients hospitalized over the same period.

**Table 1:** the number of patients (>18 years old) hospitalized annually for suicide attempts in VigilanS Regions.

|  | Nord-Pas-de-Calais (01/2015) | Basse Normandie (03/2016) | Jura department (04/2016) | Languedoc-Roussillon (05-06/2016) | Bretagne (06/2016) | Haute Normandie (03/2017) |
| --- | --- | --- | --- | --- | --- | --- |
| 2012 | 7000 | 2572 | 492 | 2105 | 5325 | 3200 |
| 2013 | 6787 | 2307 | 409 | 1565 | 5242 | 3103 |
| 2014 | 7298 | 2221 | 331 | 2357 | 4998 | 2703 |
| 2015 | 7133 | 2028 | 353 | 2372 | 4716 | 2578 |
| 2016 | 6922 | 2017 | 310 | 2441 | 4394 | 2524 |
| 2017 | 7045 | 1967 | 363 | 2291 | 4598 | 2488 |
| 2018 | 7277 | 1887 | 353 | 2273 | 4718 | 2539 |
| 2019 | 7256 | 1933 | 311 | 2006 | 4346 | 2442 |
| 2020 | 6440 | 1705 | 277 | 1972 | 4214 | 2143 |
| 2021 | 6604 | 1777 | 303 | 1879 | 4111 | 2239 |
| 2022 | 6394 | 1832 | 277 | 2422 | 4294 | 2096 |
| Total | 76156 | 22246 | 3779 | 23683 | 50956 | 28055 |

**Table 2:** cox proportional hazards model.

| Region | Variable | coef | HR | 95%CI |  | se(coef) | z | p |
| --- | --- | --- | --- | --- | --- | --- | --- | --- |
|  |  |  |  | lower | upper |  |  |  |
| Nord-Pas-de-Calais | Vigilans | -0,423 | 0,655 | 0,63 | 0,681 | 0,02 | -21,287 | <0,0001 |
|  | sexe(male) | 0,019 | 1,019 | 0,983 | 1,057 | 0,018 | 1,046 | 0,296 |
|  | age | -0,005 | 0,995 | 0,994 | 0,996 | 0,001 | -8,476 | <0,0001 |
| Basse Normandie | VigilanS | -0,344 | 0,709 | 0,663 | 0,759 | 0,034 | -9,979 | <0,0001 |
|  | sexe(male) | 0,061 | 1,063 | 0,995 | 1,135 | 0,033 | 1,822 | 0,069 |
|  | age | -0,005 | 0,995 | 0,993 | 0,997 | 0,001 | -5,157 | <0,0001 |
| Jura | Vigilans | -0,244 | 0,783 | 0,664 | 0,924 | 0,084 | -2,901 | <0,01 |
|  | Sexe(male) | 0,105 | 1,11 | 0,944 | 1,306 | 0,083 | 1,266 | 0,206 |
|  | age | -0,006 | 0,994 | 0,989 | 0,999 | 0,003 | -2,181 | <0,05 |
| Languedoc-Roussillon | VigilanS | -0,207 | 0,813 | 0,759 | 0,871 | 0,035 | -5,852 | <0,0001 |
|  | sexe(male) | 0,139 | 1,149 | 1,072 | 1,231 | 0,035 | 3,928 | <0,0001 |
|  | age | -0,009 | 0,991 | 0,989 | 0,993 | 0,001 | -8,889 | <0,0001 |
| Bretagne | VigilanS | -0,376 | 0,687 | 0,658 | 0,717 | 0,022 | -17,151 | <0,0001 |
|  | sexe | 0,098 | 1,103 | 1,058 | 1,149 | 0,021 | 4,629 | <0,0001 |
|  | age | -0,008 | 0,992 | 0,991 | 0,994 | 0,001 | -11,744 | <0,0001 |
| Haute Normandie | VigilanS | -0,381 | 0,683 | 0,642 | 0,727 | 0,032 | -11,978 | <0,0001 |
|  | sexe(male) | 0,051 | 1,052 | 0,993 | 1,114 | 0,029 | 1,725 | 0,085 |
|  | age | -0,005 | 0,994 | 0,993 | 0,996 | 0,001 | -6,258 | <0,0001 |

Analyzing the annual data for each region, we can observe fluctuations in the number of SA cases over time. In the Languedoc-Roussillon region, there was a marginal increase, from 2,105 cases in 2012 to 2,422 in 2022, indicating a gradual escalation in SA incidents. Conversely, the Basse Normandie region demonstrated relative stability, with minor fluctuations ranging from 1,705 cases in 2020 to 2,572 in 2012, suggesting a consistent rate of SA occurrences. Notably, several regions experienced a decrease in SA cases. Nord-Pas-de-Calais marked a significant reduction, from 7,277 cases in 2018 to 6,333 in 2022, reflecting a substantial downward trend. The Jura department showed a gradual decline, from 492 cases in 2012 to 277 in 2022, indicating a steady diminution. Similarly, Bretagne and Haute Normandie revealed continuous decreasing trends, with Bretagne’s cases declining from 5,325 in 2012 to 4,111 in 2021, and Haute Normandie’s from 3,200 in 2012 to 2,096 in 2022. This analysis elucidates the year-over-year variations in SA cases, providing a clearer understanding of the temporal dynamics and regional variations in these incidents.

### Survival analysis

#### Kaplan-Meier estimate

The Kaplan-Meier estimate, as illustrated in the figure 1, provides insights into the differences in the time to SA reiteration distribution for patients before and after VigilanSimplementation. The log-rank test revealed that these differences were statistically significant in all regions (P<0.0001).

**Fig 1.**
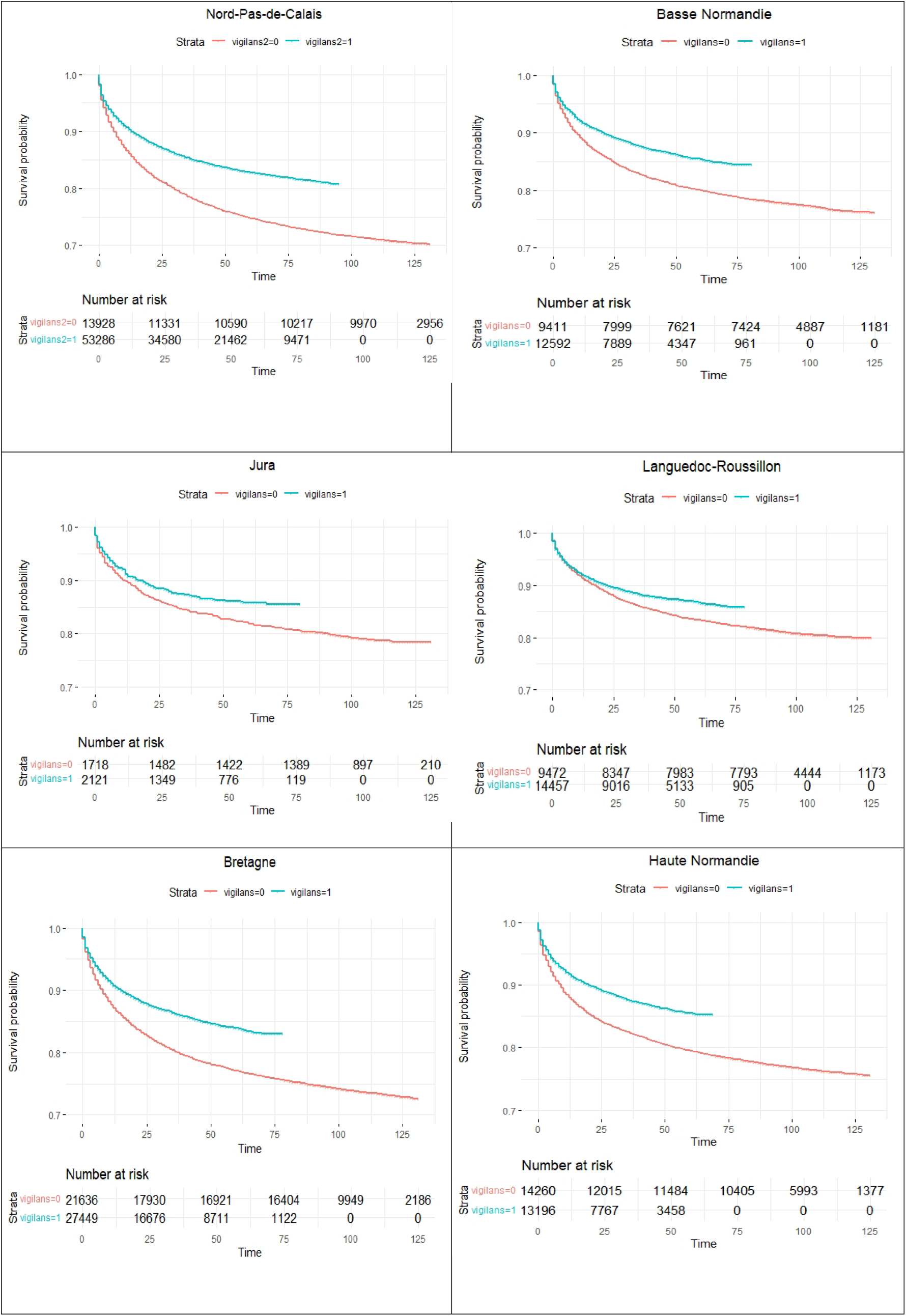
The Kaplan-Meier Survival Analysis of the number of inpatients hospitalized for suicide attempts in the PMSI system from six regions in France (Nord-Pas-de-Calais, Basse Normandie, Jura department, Languedoc-Roussillon, Bretagne and Haute Normandie), divided by before and after VigilanS project implementation, where blue is after VigilanS project implementation and red is before VigilanS project implementation.

#### Cox proportional hazards model

The Cox regressions indicate that the risk of suicide reattempt was significantly lower in all regions after VigilanS implementation. Age had a consistent negative association with this risk, meaning that as age increased, the risk decreased. Sex showed significant associations only in Languedoc-Roussillon and Bretagne, with a higher risk for female compared to male.

In the Nord-Pas-de-Calais region, “VigilanS” group has a hazard ratio (HR) of 0.655 (95% CI: 0.63 - 0.681) compared to the group without VigilanS. This means that the risk of experiencing a suicide reattempt is reduced by roughly one third when VigilanS kicks in. Gender showed no significant association with the risk of SA reattempt, while age exhibited a significant relationship: for every year increase in age, the risk of reattempt decreased by 0.5% (HR=0.995, 95% CI: 0.994 - 0.996).

Similarly, in the Basse Normandie region and the Jura department, the VigilanS group had a hazard ratio (HR) of 0.709 (95% CI: 0.663 to 0.759) and 0.783 (95% CI: 0.664 to 0.924) respectively. Moving to the Languedoc-Roussillon and Bretagne region, the VigilanS group had a hazard ratio (HR) of 0.813 (95% CI: 0.759 to 0.871) and 0.687 (95% CI: 0.658 to 0.717). Finally, in the Haute Normandie region, the VigilanS group had a hazard ratio (HR) of 0.683 (95% CI: 0.642 to 0.727)In the Haute Normandie region, a significant relationship was identified between age and the risk of reiterating suicidal attempts. However, it is crucial to differentiate the effects of age from the primary objective of this study, which is to evaluate the effectiveness of the VigilanS intervention. The VigilanS group exhibited a Hazard Ratio (HR) of 0.683 (95% CI: 0.642 to 0.727), signifying a 31.7% lower risk of suicide reattempt compared to the reference group, independent of age and gender. This HR value is of particular importance as it directly measures the impact of the VigilanS intervention, underscoring its effectiveness in reducing the risk of reattempted suicide. This highlights the effectiveness of VigilanS in mitigating suicide reattempt risk, asserting its impact at a constant age and gender basis.

## Discussion

This study critically evaluates the impact of the VigilanS project on reducing suicide attempts (SA) among patients aged 18 years and older in general hospitals throughout France. The results indicate varied outcomes: while there is a general trend of reduced SA hospitalizations per year, extended time intervals between initial and subsequent SAs, and a lowered risk of a secondary SA following an initial attempt, these trends are not uniform across all regions. In some of the six French regions where VigilanS was implemented, there have been instances of increasing SA cases. This complexity highlights that while VigilanS demonstrates potential as an effective suicide prevention tool, its impact is not universally consistent across different regions. These findings suggest the need for a region-specific approach in the implementation of Brief Contact Interventions (BCIs) in general hospitals, tailoring strategies to effectively address local dynamics in suicide prevention.Before VigilanS project, as early as 2014, one clinical trial project AlgoS examined the efficacy of BCI for suicide prevention in 23 French hospitals[22,23]. The intervention included crisis cards for first-time attempters and phone calls for multi-attempters. AlgoS served as a foundation for the development of VigilanS, an enhanced version or evolution of the intervention.Since 2015, the VigilanS project has been implemented in the region of Nord-Pas-De-Calais in France[19]. The study evaluated the effectiveness of VigilanS in reducing suicide reattempt (SR) among patients hospitalized for SA has demonstrated that the exposed group had a 5.2% SR rate compared to 22.2% in the nonexposed group, indicating its effectiveness in reducing suicide reattempt. However, the project only benefits a portion of the patients in two departments of northern France and could be extended to help more patients[16].

To date, there is limited research examining the effectiveness of Brief Contact Interventions (BCIs) in the general population and real-world conditions. Most studies on BCIs have been conducted within specific contexts, such as RCTs or targeted populations[9,10,24]. The application of BCIs in broader settings, considering factors like diverse demographics, healthcare systems, and individual circumstances, has not been extensively explored. As a result, our current understanding of the efficacy of BCIs in real-world scenarios is limited.

The number of patients hospitalized for SA in the first six regions under scope highlights the high burden of SA in these regions. These findings underscore the importance of targeted interventions and the implementation of programs like VigilanS to address the specific needs of each region for suicidal attempts incidence reductionof. The six regions where the VigilanS project was implemented show varying degrees of effectiveness. The most to least favorable outcomes of VigilanS are observed in the following regions: Nord-Pas-de-Calais, Basse Normandie, Bretagne, Haute Normandie, Jura, and Languedoc-Roussillon. The variation in outcomes underscores the necessity of region-specific interventions and highlights the importance of adapting programs like VigilanS to meet the unique challenges and needs of each region in reducing the incidence of suicide attemptsSeveral factors may explain these differences in effectiveness. Firstly, the regions had different start dates for implementing VigilanS. For instance, the Nord-Pas-de-Calais region, where VigilanS was created, began implementing the program as early as 2015, while other regions followed at later dates. These temporal differences can impact the results for each region. Furthermore, healthcare infrastructures and regional healthcare organization in France can vary across regions. Each region operates under the authority of a Regional Health Agency (Agence Régionale de Santé), which can lead to different adaptations in the implementation of VigilanS. These contextual differences can influence the results for each regionThe observed improvement in SA-free time highlights the importance of timely and targeted interventions offered by the VigilanS project. By providing support and resources to individuals at risk of reattempting suicide, the intervention has been successful in improving patient outcomes. The positive impact of the VigilanS project emphasizes the importance of implementation of similar initiatives aimed at preventing suicide reattempts.

### Strengths of the Study

1. Real-World Evidence (RWE): The study provides valuable real-world evidence by evaluating the effectiveness of the VigilanS intervention in actual clinical practice and real-life conditions. This approach enhances the relevance and applicability of the findings to everyday healthcare settings, offering insights into the intervention’s impact in naturalistic settings.
2. Large Population Size: The significant population size of the study increases its statistical power and precision. With a large sample, the study is more capable of detecting meaningful effects and drawing reliable conclusions, thereby enhancing the robustness of its findings.
3. Extensive Study Period: The study covers a comprehensive period of 11 years, which includes 5 to 7 years post-VigilanS implementation. This extended duration allows for a thorough analysis of the long-term effects and sustainability of the VigilanS intervention, providing a deeper understanding of its impact over time.

### Limitations of the Study

1. Unnoticed or unrecognized SAs: some suicide attempts may go unnoticed or unrecognized by clinicians, resulting in underreporting and the exclusion of such cases from the analysis. These attempts may not be coded as suicide attempts in the PMSI system, leading to an underestimation of the true phenomenon.
2. Incomplete capture of suicide attempts: The study relied on the PMSI system, that do not capture all suicide attempts. Non-hospitalized attempts includes those treated in primary care settings, cases without healthcare involvement (spontaneous recovery, SA resulting in death) [25]. Therefore, the study’s findings may not fully reflect the effectiveness of BCIs.
3. In this study, VigilanS was evaluated in the first six regions of France that adopted it. Therefore, our results may not apply to all regions of the country. Evaluation in the other regions is necessary to assess the project’s effectiveness on a broader scale, and to determine if similar outcomes can be achieved in other settings.
4. Potential bias due to missing data: Some patient’s data from emergency departments were not present in the PMSI database, it could introduce bias into the final results.
5. Mixture of included and non-included patients: after the implementation of VigilanS, the intervention was not offered overnight to all patients, resulting in a mixture of included and non-included patients in our “post-VigilanS” group. Despite this limitation, the study still elicited an effect. Ideally, it would have been interesting to identify patients who were actually included in the intervention, but this process would have been complex and resource-intensive[26]. Therefore, the study’s findings represent a pragmatic evaluation of VigilanS, under real-life circumstances of an incomplete coverage of all eligible patients.

## Conclusion

The study provides evidence that implementing the VigilanS project following suicide attempts in general hospitals can be an effective approach to reducing suicide rates in France. These findings are particularly important given the high suicide rate in France and the need for effective suicide prevention strategies. By using BCIs to monitor and intervene with patients at high risk of suicide, the VigilanS project may offer a valuable prevention tool for healthcare stakeholders. The study’s results have implications for suicide prevention efforts in other countries and highlight the potential benefits of interventions into standard care for suicide prevention.

## Declarations

### Ethics Approval and Consent to Participate

The Commission Nationale de l’Informatique et des Libertés (CNIL), the national data protection authority of France, waived ethical approval for this work. The study used fully anonymized administrative hospitalization data from the French PMSI-MCO database. According to French law and CNIL regulations, secondary analyses of anonymized administrative health data do not require approval from an Institutional Review Board or written informed consent from patients.

### Consent for Publication

Not applicable. This study used anonymized administrative data that contain no identifiable information.

### Data Availability

The data used in this study originate from the French national PMSI-MCO hospital discharge database. Due to legal restrictions imposed by CNIL and the Agence Technique de l’Information sur l’Hospitalisation (ATIH), raw PMSI data cannot be publicly shared. Aggregated data supporting the findings of this work are available from the corresponding author upon reasonable request. Researchers wishing to access PMSI data must submit an application through ATIH and follow national authorization procedures.

### Author Contributions

Xavier Xu WANG: Conceptualization, data extraction, data curation, statistical analysis, interpretation, manuscript drafting, manuscript revision.

## Acknowledgements

The author thanks the VigilanS program team for their contribution to the development and implementation of the national intervention, in particular Antoine Messiah and Guillaume Vaiva, whose work in the program’s operational deployment has contributed to the broader context in which this analysis was conducted. The author also acknowledges ATIH for providing access to PMSI data under national authorization procedures. All analyses, interpretations, and conclusions presented in this manuscript are solely those of the author.

## Appendix 1

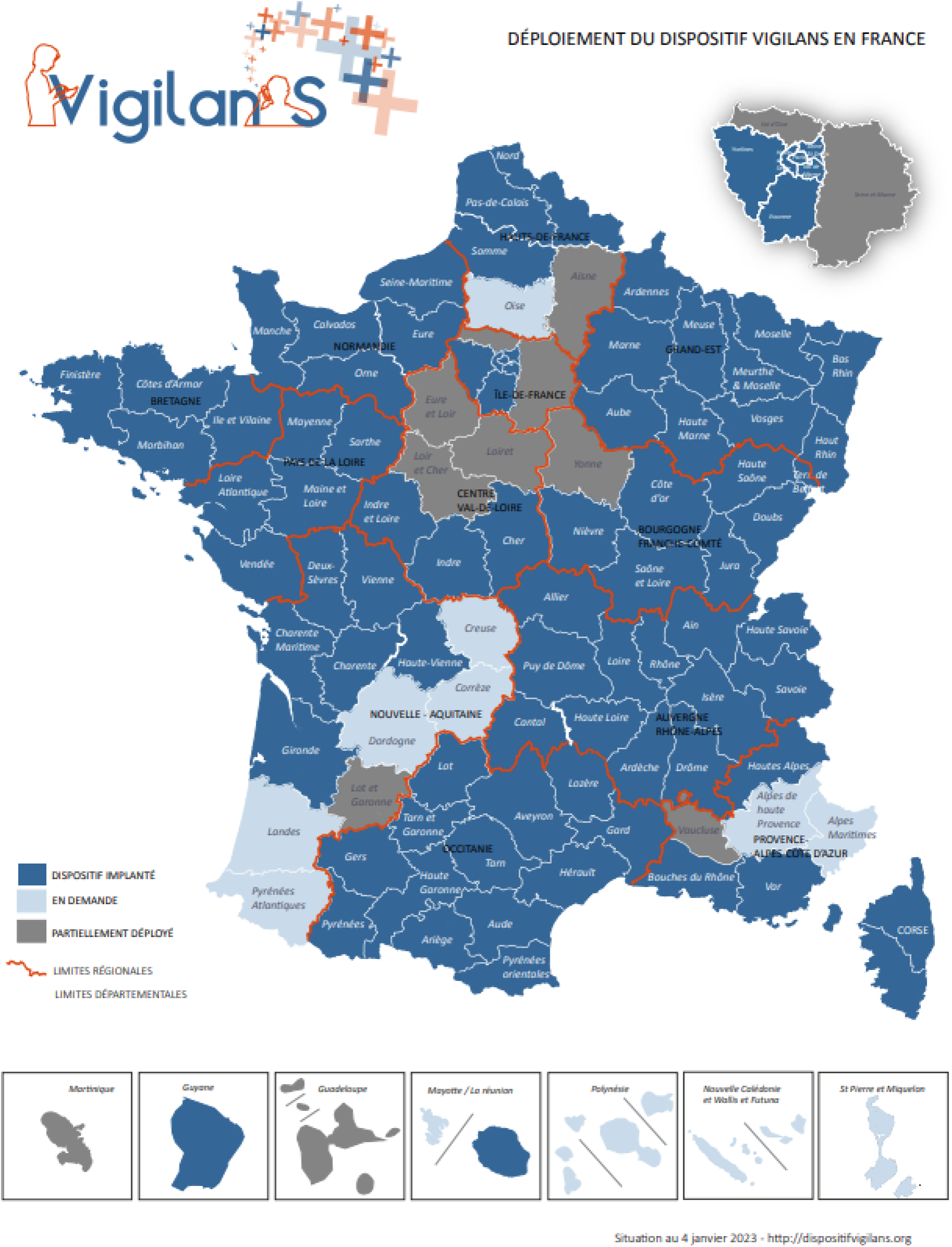

## Reference

1. Pompili M, O’Connor RC, van Heeringen K. Suicide Prevention in the European Region. Crisis. 1 mars 2020;41(Supplement 1):S8⍰20.

2. Hawton K, Witt KG, Salisbury TLT, Arensman E, Gunnell D, Hazell P, et al. Psychosocial interventions following self-harm in adults: a systematic review and meta-analysis. Lancet Psychiatry.août 2016;3(8):740⍰50.

3. Riblet NBV, Shiner B, Young-Xu Y, Watts BV. Strategies to prevent death by suicide: meta-analysis of randomised controlled trials. Br J Psychiatry. 2017;210(6):396⍰402.

4. du Roscoat E, Legleye S, Guignard R, Husky M, Beck F. Risk factors for suicide attempts and hospitalizations in a sample of 39,542 French adolescents. J Affect Disord. 15 janv 2016;190:517⍰21.

5. Inagaki M, Kawashima Y, Kawanishi C, Yonemoto N, Sugimoto T, Furuno T, et al. Interventions to prevent repeat suicidal behavior in patients admitted to an emergency department for a suicide attempt: a meta-analysis. J Affect Disord. 2015;175:66⍰78.

6. Martínez-Alés G, Cruz Rodríguez JB, Lázaro P, Domingo-Relloso A, Barrigón ML, Angora R, et al. Cost-effectiveness of a Contact Intervention and a Psychotherapeutic Program for Post-discharge Suicide Prevention. Can J Psychiatry. août 2021;66(8):737⍰46.

7. López-Goñi JJ, Goñi-Sarriés A. Effectiveness of a telephone prevention programme on the recurrence of suicidal behaviour. One-year follow-up. Psychiatry Res. août 2021;302:114029.

8. Amadéo S, Rereao M, Malogne A, Favro P, Nguyen NL, Jehel L, et al. Testing Brief Intervention and Phone Contact among Subjects with Suicidal Behavior: A Randomized Controlled Trial in French Polynesia in the Frames of the World Health Organization/Suicide Trends in At-Risk Territories Study. Ment Illn [Internet]. 2015 [cité 16 août 2016];7(2). Disponible sur: http://www.ncbi.nlm.nih.gov/pmc/articles/PMC4620282/

9. Tay JL, Li Z. Brief contact interventions to reduce suicide among discharged patients with mental health disorders-A meta-analysis of RCTs. Suicide Life Threat Behav. déc 2022;52(6):1074⍰95.

10. Doupnik SK, Rudd B, Schmutte T, Worsley D, Bowden CF, McCarthy E, et al. Association of Suicide Prevention Interventions With Subsequent Suicide Attempts, Linkage to Follow-up Care, and Depression Symptoms for Acute Care Settings: A Systematic Review and Meta-analysis. JAMA Psychiatry. 1 oct 2020;77(10):1021.

11. Duhem S, Berrouiguet S, Debien C, Ducrocq F, Demarty AL, Messiah A, et al. Combining brief contact interventions (BCI) into a decision-making algorithm to reduce suicide reattempt: the VigilanS study protocol. BMJ Open. 23 oct 2018;8(10):e022762.

12. Fossi Djembi L, Vaiva G, Debien C, Duhem S, Demarty AL, Koudou YA, et al. Changes in the number of suicide re-attempts in a French region since the inception of VigilanS, a regionwide program combining brief contact interventions (BCI). BMC Psychiatry. déc 2020;20(1):26.

13. Fossi LD, Debien C, Demarty AL, Vaiva G, Messiah A. Suicide reattempt in a population-wide brief contact intervention to prevent suicide attempts: The VigilanS program, France. Eur Psychiatry. 2021;64(1):e57.

14. Chan Chee C, Paget LM. Le Recueil d’information médicalisé en psychiatrie (RIM-P) : un outil nécessaire pour la surveillance des hospitalisations suite à une tentative de suicide. Rev DÉpidémiologie Santé Publique. 1 sept 2017;65(5):349⍰59.

15. Plancke L, Ducrocq F, Clément G, Chaud P, Haeghebaert S, Amariei A, et al. Les sources d’information sur les tentatives de suicide dans le Nord - Pas-de-Calais. Apports et limites. Rev DÉpidémiologie Santé Publique. déc 2014;62(6):351⍰60.

16. Plancke L, Amariei A, Danel T, Debien C, Duhem S, Notredame CE, et al. Effectiveness of a French Program to Prevent Suicide Reattempt (VigilanS). Arch Suicide Res. 5 mars 2020;1⍰12.

17. Domin JP. Le Programme de médicalisation des systèmes d’information (PMSI): De l’indicateur de comptabilité hospitalière au mode de tarification (1982-2012). Hist Médecine Santé. 1 nov 2013;(4):69⍰87.

18. Silverman MM, Berman AL, Sanddal ND, O’carroll PW, Joiner TE. Rebuilding the tower of babel: a revised nomenclature for the study of suicide and suicidal behaviors part 1: background, rationale, and methodology. Suicide Life Threat Behav. 2007;37(3):248⍰63.

19. Vaiva G, Plancke L, Amariei A, Demarty AL, Lardinois M, Creton A, et al. Évolutions du nombre de tentatives de suicide dans le Nord-PasdeCalais depuis l’implantation de VigilanSlll: premières estimations. L’Encéphale. janv 2019;45:S22⍰6.

20. Ministère des affaires sociales et de la santé. CIM-10 à usage PMSI - Classification statistiques internationale des maladies et des problèmes de santé connexes. 2018; Disponible sur: https://www.atih.sante.fr/sites/default/files/public/content/3295/cim-10fr_2018_v1_provisoire.pdf

21. George B, Seals S, Aban I. Survival analysis and regression models. J Nucl Cardiol Off Publ Am Soc Nucl Cardiol. août 2014;21(4):686⍰94.

22. Messiah A, Notredame CE, Demarty AL, Duhem S, Vaiva G, Investigators on behalf of A. Combining green cards, telephone calls and postcards into an intervention algorithm to reduce suicide reattempt (AlgoS): P-hoc analyses of an inconclusive randomized controlled trial. PLOS ONE. févr 2019;14(2):e0210778.

23. Demesmaeker A, Chazard E, Vaiva G, Amad A. Risk Factors for Reattempt and Suicide Within 6 Months After an Attempt in the French ALGOS Cohort: A Survival Tree Analysis. J Clin Psychiatry. 18 févr 2021;82(1):20m13589.

24. Lin Y, Liu S, Chen S, Sun F, Huang H, Huang C, et al. Brief Cognitive-based Psychosocial Intervention and Case Management for Suicide Attempters Discharged from the Emergency Department in Taipei, Taiwan: A Randomized Controlled Study. Suicide Life Threat Behav. juin 2020;50(3):688⍰705.

25. Jollant F, Hawton K, Vaiva G, Chan-Chee C, du Roscoat E, Leon C. Non-presentation at hospital following a suicide attempt: a national survey. Psychol Med. mars 2022;52(4):707⍰14.

26. Broussouloux Sandrine, Gallien Yves, Fouillet Anne, Mertens Clément. Évaluation d’efficacité de VigilanS de 2015 à 2017, dispositif de prévention de la réitération suicidaire [Internet]. 2023 sept p. 8. Disponible sur: https://www.santepubliquefrance.fr/maladies-et-traumatismes/sante-mentale/suicides-et-tentatives-de-suicide/documents/enquetes-etudes/evaluation-d-efficacite-de-vigilans-de-2015-a-2017-dispositif-de-prevention-de-la-reiteration-suicidaire

